# Insights into Essential Tremor and Essential Tremor-Plus from Common Variants

**DOI:** 10.64898/2025.12.02.25341457

**Authors:** Miranda Medeiros, Dylan Gharibian, Patrick A. Dion, Guy A. Rouleau

## Abstract

**Intro:** The heterogeneity of Essential Tremor (ET) complicates how it is diagnosed and studied. ET-Plus is a concept proposed to help explain the overlap of ET clinical features with soft signs of other neurological disorders. We aimed to better understand ET by comparing brain maps informed by ET common variant to those derived from phenotypes involved in ET-Plus. A further goal was to enhance the diagnostic precision of ET by accounting for shared neurobiological signals between ET and ET-Plus–related phenotypes.

**Methods:** Phenotype variant association mapping to the brain was done for ET, Parkinson’s disease (PD), dystonia, and cognition across adult mouse whole brain and cerebellum spatial transcriptomic data through gsMap. Separately, ET genome-wide association study (GWAS) summary statistics were conditioned on PD and cognition to account for shared genetic signals. Using both raw and conditioned GWASes, ET polygenic risk scores (PRS) were calculated across patient cohorts and controls, and their respective ability to classify ET at the 90th percentile was compared using McNemar’s test.

**Results:** Spatial mappings of GWAS signals revealed many shared associations between phenotypes. The raw ET PRS model preformed 1.33% (95% CI: [0.30% - 2.35%]; p = 0.0129) better than the conditioned ET PRS model.

**Conclusion:** We lack the ability to decern ET from phenotypes involved in ET-Plus using existing common variant disease associations. Efforts to isolate a core genetic signal for ET by de-noising shared associations reduced the accuracy of patient classification. A more effective strategy to studying ET may be to leverage its heterogeneity rather than attempt to isolate it.

## Introduction

Essential Tremor (ET) is a complex condition and the most common movement disorder globally with a prevalence that increases with age. ET is defined as an isolated action tremor present in bilateral upper extremities for at least three years. ET tremors may or may not also be present in the head, voice, or lower limbs. However, clinical presentation is varied given the heterogenous symptoms observed across cases.^1^ This in-part lends itself to the high rate of misdiagnosis of ET as other movement disorders such as Parkinson’s disease (PD).^2^

The heterogeneity of ET incited the creation of the “Essential Tremor Plus” phenotype (ET-Plus), where “Pure ET” does not allow for any signs of other neurologic disease, the ET-Plus classification does.^1^ These signs of other neurologic disease, also known as “soft signs” may include difficulties in tandem gait, dystonic posturing, rest tremor, and mild memory impairment among other mild neurological signs. Defining ET in a patient using this classification method becomes increasingly complex as the cut-off of what constitutes a soft sign is not objectively defined. In the clinic soft signs are frequently observed, so much so that it has been proposed that ET-Plus is clinically more common than Pure-ET.^3^

There is a growing notion that ET is not monosymptomatic but rather is a heterogeneous disorder, likely representing a broad disease spectrum encompassing many different possible symptoms and features. It is not unreasonable to suspect that ET may not just be an isolated action tremor, as many other progressive later-onset movement disorders such as Parkinson’s disease or Huntington’s disease are accompanied by other motor and not-motor features. The progressive nature of ET also adds complexity to the notion of ET-Plus, since additional symptoms may begin to present the longer a patient suffers from ET. As such, it is unclear if Pure-ET may be a snapshot of the disease before it has sufficiently progressed into a potential polysymptomatic disorder captured by the ET-Plus definition. In other words, whether Pure-ET or ET-Plus are subtypes of ET, or normal stages in the progression of ET is unknown.^4^

Given that ET progresses over time, a proposed ET model that ties into the ET-Plus concept suggests that ET begins as Pure-ET and may then evolve into ETc (for cerebellar involvement, i.e. ataxic symptoms), ETp (for mild parkinsonism), ETd (for mild dystonia), and ETcog (for cognitive impairment).^3^ Over time, these soft signs belonging to distinct ET-Plus subtypes may evolve into overt parkinsonism, cerebellar ataxia, dystonia, or dementia respectively with antecedent ET.^3^ With the ET-Plus subtypes of this model as our basis, we aimed to understand how common ET genetic risk compared to the genetic risk of PD, ataxia, dystonia, and cognitive impairment (representing ET-Plus related phenotypes) spatially affect the brain. We also aimed to define a de-noised genetic profile of ET to improve diagnosis by controlling for the genetic risk of ET-Plus related phenotypes (PD, ataxia, dystonia, and cognitive impairment).^3^ As ET-Plus appears to be the dominant form of ET, it is likely that ET genetic studies have suffered from noise attributed to a genetic vulnerability overlap with related neurological disorders. As such, resolving the genetic underpinning of ET has remained elusive. Overall, we aim to identify a potential genetic core of ET which may allow for improved cohort selection for studies which may help to elucidate disease mechanisms and treatments.

## Methods

### Spatial differences between ET and ET-Plus phenotypes

To reveal associations between spatial transcriptomics and phenotype specific genetic risk of ET and ET-Plus related phenotypes, we used gsMap.^5^ GsMap is a tool which allows for the pairing of spatial transcriptomics and genome wide association study (GWAS) summary statistics to resolve which regions or cells of a tissue are enriched for disease associated single nucleotide polymorphisms (SNPs). In other word, the tool resolves the spatial mapping of complex trait-associated cells.^5^

#### SNP phenotype associations from GWASes

To identify relevant phenotype associated cells and regions, full summary statistics from large and current GWASes were gathered for ET, PD, dystonia, and cognition phenotypes respectively. Full summary statistics contain the SNP-phenotype associations needed for gsMap.^5^ ET summary statistics are taken from Skuladottir *et. al*,^6^ those for PD were taken from Nalls *et. al*,^7^ those for dystonia came from Laabs *et. al*,^8^ and those for cognition were sourced from Li *et. al*.^9^ Ataxia was a considered phenotype since it tends to show soft signs in ET-Plus,^3^ however the genetic drivers of ataxia tend to be repeat expansions or follow autosomal dominant inheritance patterns.^10^ Despite this, there does exist an ataxia GWAS, but it represents the Machado-Joseph form of the disease which is a rare condition and not very informative to examine in the context of ET.^11^ As such, ataxia was excluded from the study since ET relevant ataxia is not well characterized by GWAS and thus would be ill-suited for gsMap analysis. Additionally, it is worth noting that though the dystonia GWAS lacks power compared to the other included phenotypes, it was still examined as gsMap analyses run independently and the selected GWAS offered the most available power being the largest and most current dystonia GWAS. The characteristics of the selected GWASes are described by esupp Table S1.

#### Spatial datasets

Based on available public datasets, we opted to identify brain region-trait associations across the mouse brain. Spatial transcriptomic data covering the whole adult mouse brain was taken from Chen *et. al*.^12^ Single cell annotations were transferred to the spatial dataset through scVI integration from the scvi-tools package.^13^ To study the cerebellum, mouse cerebellum spatial transcriptomic data with single cell annotations from Hao *et. al*,^14^ was obtained.

#### Brain mapping with gsMap

The gsMap pipeline was followed as outlined in the Yang lab guide.^5^ The first step consisted of learning latent representations for spots across spatial datasets using *run_find_latent_representations*. Next, using *run_latent_to_*gene homogeneous spots were identified, homologous gene transformation was done to convert mouse genes to human equivalents for later GWAS integration, and gene specificity scores (GSS) were assigned to each spot. After this, GSS were linked to SNPs and stratified LD scores were computed using *run_generate_ldscore* for each chromosome. Specifically, GSS were assigned to SNPs using transcription start site (TSS) proximity and enhancer-gene linking. Reference data as well as homologous gene transformation list were obtained from gsMap’s resource bundle.^5^ Finally, GWAS summary statistics for ET, PD, dystonia, and cognition were used to associate spots with traits through spatial LDSC using *run_spatial_ldsc*. This was done separately for the whole mouse brain and the mouse cerebellum for all GWAS summary statistics (ET, PD, dystonia, cognition; GWASes detailed in esupp Table S1).

Spatially mapped trait associations were then analysed using Cauchy combination which aggregated p-values of individual spots to identify which cell types and brain regions were most strongly associated with a given phenotype using *run_cauchy_combination*. Then final trait spatial association reports, including visualizations across spatial data, were generated using *run_report*. P-Cauchy values were corrected for multiple hypothesis testing using the Bonferroni method, where for the whole mouse brain the p-value significance threshold = 0.05/(48 annotations × 4 phenotypes) = 2.60×10^-4^, and for the cerebellum the p-value significance threshold = 0.05/(22 annotations × 4 phenotypes) = 5.68×10^-3^.

### De-noised ET GWAS and Polygenic Risk Scoring

To attempt to eliminate some of the noise attributed to the heterogeneity of ET and to identify a unique genetic profile for the condition, we adjusted ET GWAS associations based on shared ET-Plus related phenotype genetic risk. This was done with the aim of improving ET polygenic risk scoring to better discriminate ET samples from non-ET individuals (PD, dystonia, cerebellar ataxia, cognitively impaired, and health samples).

#### Conditioning Essential Tremor on ET-Plus related phenotypes

The mtCOJO^15^ tool from the GCTA^16^ suite was used to condition the ET GWAS summary statistics on the GWAS summary statistics of ET-Plus related phenotypes (PD and cognition; esupp Table 1). The dystonia GWAS summary statistics had to be removed from the analysis, due to its lack of power compared to the other GWASes, meaning that the ET summary statistics were conditioned on PD and cognition summary statistics. 1000 Genome^17^ European linkage disequilibrium reference data was used as reference for the multi-trait GWAS conditioning. SNPs that were deemed problematic due to large differences in effect allele frequencies between phenotypes were removed to avoid spurious associations due to ambiguous SNPs. A Manhattan plot of conditioned ET GWAS was then constructed in R using ggplot (esupp Figure S1).

#### Essential Tremor Polygenic Risk Scoring

A cohort of whole genome sequenced ET patients, PD patients, dystonia patients, cerebellar ataxia patients, and unaffected neurologically normal controls all above the age of 65 years old (CDR date >= 65) were assembled through the All of Us researcher workbench.^18^ An age cutoff of 65 years old was selected to attempt to minimize the number of samples who may eventually develop ET but had yet to do so. This age was chosen since ET has a bimodal age of onset, where most patients develop ET later in life, past their 4^th^ decade,^19^ and the prevalence of ET is well characterized in the 65 years of age and over population as roughly 5%.^20^

Quality control at the sample level consisted of retaining only samples with European inferred genetic ancestry as predicted by All of Us^18^ and removing related samples (second degree relatedness or closer) using the KING algorithm^21^. Additionally, if a sample had an ambiguous movement disorder diagnosis (i.e. was diagnosed with more than one movement disorder among ET, PD, dystonia, or ataxia) they were removed from the cohort. Cognitively impaired samples that carried any movement disorder diagnosis were also removed. Healthy individuals were selected as those who carried no diagnosis. Due to the large number of healthy samples and cognitively impaired samples compared to ET, PD, dystonia, and cerebellar ataxia patient cohorts, a random subset of 1,000 cognitively impaired patients above the age of 65 years old and a random subset of 2,000 healthy individuals were select prior to quality control. Following quality control, the final cohort consisted of 4,377 samples described in esupp Table S2.

Variant quality control involved, briefly, the removal of: SNPs with a minor allele frequency of < 1%, SNPs with a genotyping missingness rate of > 1%, and SNPs failing Hardy-Weinberg Equilibrium (1×10^-5^). Ambiguous SNPs were identified (palindromic SNPs with effect allele frequencies between 0.4 and 0.6) and removed. Additionally, only biallelic and autosomal SNPs were retained for analysis. Full summary statistics from the most recent and largest ET GWAS^6^ available was used to calculate native ET SNP posterior effect estimates using PRS-CS.^22^ Individual level native ET polygenic risk scores (PRS) were generated for all samples in the assembled cohorts using PLINK^23^ to sum SNP posterior estimates per genotype. PRS scores were then corrected by regression out the top 10 genetic principal components.

The conditioned ET PRS was then calculated using the same methodology for all samples in the cohort but using the ET-Plus phenotype adjusted GWAS summary statistics generated by mtCOJO rather than the direct summary statistics taken from the ET GWAS^6^.

#### Essential Tremor Polygenic Risk Score Model Performance

The ability to discriminate ET patients from non-ET samples among the individuals in the highest PRS groups was used to evaluate the performance of the conditioned ET PRS compared to native unadjusted ET PRS model. This was done through diagnostic test evaluations for sensitivity, specificity, positive likelihood ratio, negative likelihood ratio, positive predictive value, negative predictive value, and accuracy using a ≥90th percentile (top percentile) cut-off of PRS for ET patient classification. Accuracy, negative predictive value, and positive predictive value were calculated with a disease prevalence of 5%.

To directly compare the two PRS models, a two-sided McNemar paired proportion test was used to evaluate the ability to correctly decern ET patients from non-ET individuals between the two PRS models at their respective 90^th^ percentile cutoffs.

## Results

### ET and ET-Plus phenotype spatially mapped GWAS signatures

Across the adult mouse whole brain and cerebellum, the spatial regions and cell types most strongly associated to a given trait as determined by Cauchy combination are described in Table 1 (top five spatial-phenotype pairs shown ranked by smallest p-value; complete results in esupp Table S3).

**Table 1:** Most significant associations across phenotypes for whole brain and cerebellum.

| *Spatial Data* | *Phenotype* | *Annotation* | *P Cauchy* |
| --- | --- | --- | --- |
| *Adult Mouse Brain* | Cognition | Cortex Layer 6 | 1.33E-18 |
| *Adult Mouse Brain* | Cognition | Subiculum | 2.73E-17 |
| *Adult Mouse Brain* | Cognition | Cortex Layer 2/3 | 1.83E-16 |
| *Adult Mouse Brain* | Cognition | Stratum lacunosum / raditum of hippocampus cornu ammonis area 1 (CA1) | 1.99E-16 |
| *Adult Mouse Brain* | Cognition | Cortex Layer 5 | 2.17E-16 |
| *Adult Mouse Brain* | Cognition | Cortex Layer 4 | 2.67E-16 |
| *Adult Mouse Cerebellum* | Cognition | Golgi cells | 1.47E-10 |
| *Adult Mouse Cerebellum* | Cognition | Oligodendrocyte (ODC) | 2.04E-10 |
| *Adult Mouse Cerebellum* | Cognition | Granular layer | 3.80E-10 |
| *Adult Mouse Cerebellum* | Cognition | Unipolar brush cells (UBC) | 3.84E-10 |
| *Adult Mouse Cerebellum* | Cognition | Granule cells | 4.61E-10 |
| *Adult Mouse Cerebellum* | Parkinson's Disease | Bergmann glia | 1.07E-06 |
| *Adult Mouse Cerebellum* | Parkinson's Disease | Purkinje layer | 1.56E-06 |
| *Adult Mouse Brain* | Essential Tremor | Cortex Layer 5 | 4.11E-06 |
| *Adult Mouse Cerebellum* | Parkinson's Disease | Purkinje cells | 6.98E-06 |
| ***Adult Mouse Cerebellum*** | **Essential Tremor** | **Oligodendrocytes (ODC)** | **1.55E-05** |
| *Adult Mouse Brain* | Parkinson's Disease | Substantia nigra<br>/ Ventral segmental area | 2.09E-05 |
| ***Adult Mouse Brain*** | **Essential Tremor** | **Subiculum** | **2.87E-05** |
| ***Adult Mouse Cerebellum*** | **Essential Tremor** | **Purkinje layer** | **6.47E-05** |
| *Adult Mouse Cerebellum* | Parkinson's Disease | Golgi cells | 7.44E-05 |
| ***Adult Mouse Cerebellum*** | **Essential Tremor** | **Golgi cells** | **9.63E-05** |
| ***Adult Mouse Cerebellum*** | **Essential Tremor** | **Bergmann glia** | **9.84E-05** |
| *Adult Mouse Brain* | **Essential Tremor** | **Hippocampus cornu ammonis area 1**<br>**(CA1)** | **1.23E-04** |
| ***Adult Mouse Cerebellum*** | **Essential Tremor** | **Purkinje cells** | **1.42E-04** |
| ***Adult Mouse Brain*** | **Essential Tremor** | **Cortex Layer 6** | **1.71E-04** |
| *Adult Mouse Cerebellum* | Parkinson's Disease | Ependymal cells | 2.35E-04 |

Note: Significant essential tremor results are denoted in bold.

Notably, across the adult mouse brain, ET and related phenotypes’ genetic risk tend to be more closely associated to brain regions rather than specific cell types. The exception is dystonia which is more strongly associated with GABAergic interneurons (p-Cauchy = 1.93e-02) rather than any specific brain region, however the dystonia findings do not survive multiple hypothesis correction.

There exists no brain region uniquely mapped by ET GWAS signatures. The most significant association between ET GWAS signals and the adult mouse brain are found in layer 5 of the cortex (p-Cauchy = 4.11e-06) whereas the most significant association between ET GWAS signals and the mouse cerebellum are found in oligodendrocytes cells (p-Cauchy = 1.55e-05). Spatial maps of GWAS signatures and how they compared between phenotypes across the whole adult mouse brain excluding the cerebellum are shown in Figure 1.

**Figure 1.**
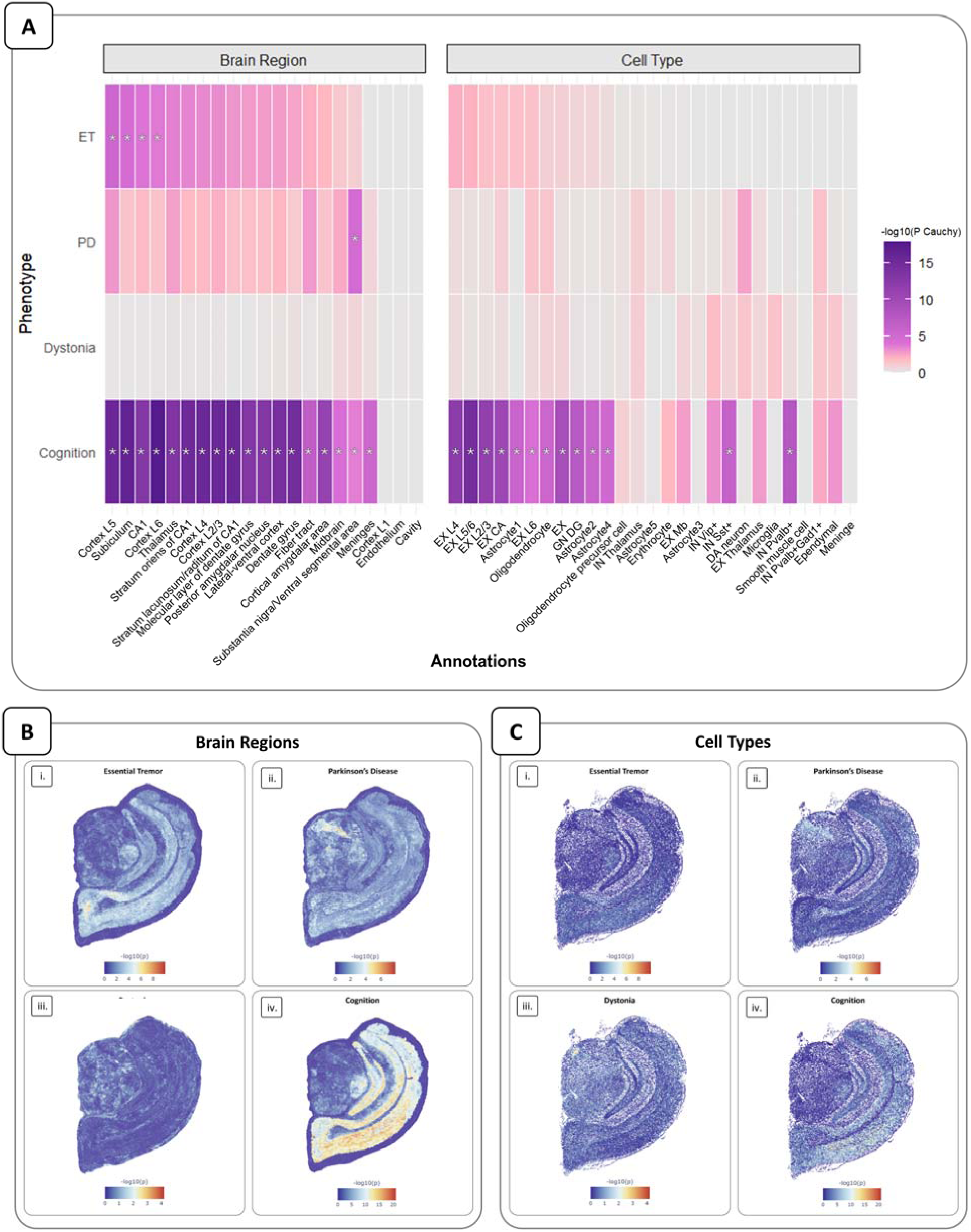
Phenotype GWAS association mapping to adult mouse whole brain. (A) Heatmap depicting Cauchy combination test aggregate p-values from gsMap GWAS informed mapping of Essential Tremor (ET), Parkinson’s disease (PD), dystonia, and cognition onto adult mouse brain spatial transcriptomics. Results are shown separate for brain regions and cell types. All annotations are ranked from what is most significantly associated with ET to least associated for brain regions and cell types respectively. Deeper shades of purple indicate smaller p-values, whereas pink represents larger p-values. Significant results are denoted with an asterisk (*) given a Bonferroni p-value threshold = 2.60×10^-4^. (B) gsMap brain region results for (i) Essential Tremor, (ii) Parkinson’s disease, (iii) dystonia, (iv) and cognition, mapped to adult mouse brain spatial transcriptomic data. Each point presents that spot’s association with trait specific GWAS signatures, with smaller Cauchy P-values in red and larger ones in blue. (C) gsMap cell type results for (i) Essential Tremor, (ii) Parkinson’s disease, (iii) dystonia, (iv) and cognition, mapped to adult mouse brain spatial transcriptomic data. Each point presents that spot’s association with trait specific GWAS signatures, with smaller Cauchy P-values in red and larger ones in blue.

The significant associations between GWAS signatures and brain spatial mapping is more extensive in the cerebellar data not just for ET but for PD and cogniton aswell. There again exists no region nor cell type uniquely associated with ET GWAS signatures. Spatial maps of GWAS signatures and how they compared between phenotypes across the whole adult mouse cerebellum is shown in Figure 2.

**Figure 2.**
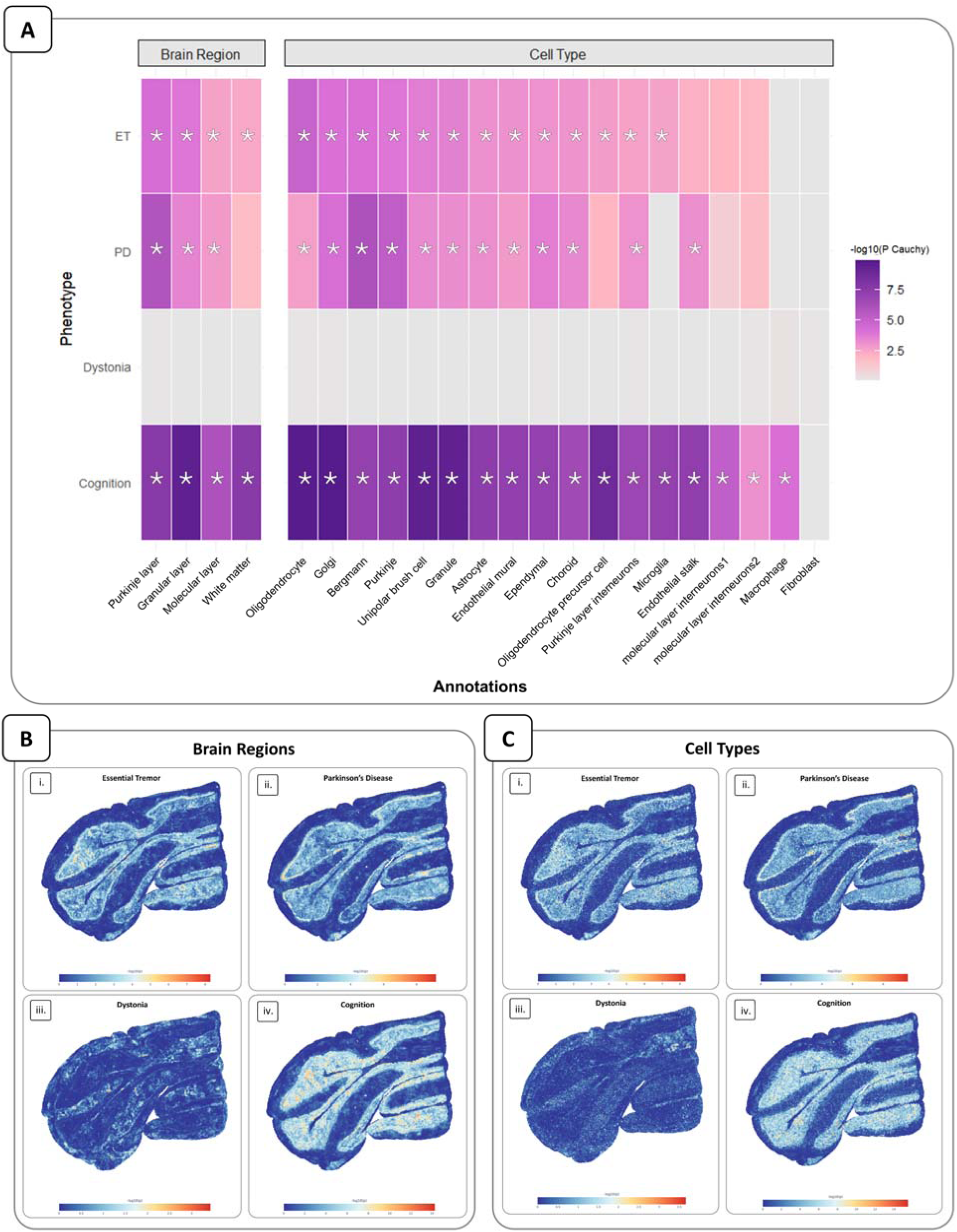
Phenotype GWAS association mapping to adult mouse cerebellum. (A) Heatmap depicting Cauchy combination test aggregate p-values from gsMap GWAS informed mapping of Essential Tremor (ET), Parkinson’s disease (PD), dystonia, and cognition onto adult mouse cerebellum spatial transcriptomics. Results are shown separate for brain regions and cell types. All annotations are ranked from what is most significantly associated with ET to least associated for brain regions and cell types respectively. Deeper shades of purple indicate smaller p-values, whereas pink represents larger p-values. Significant results are denoted with an asterisk (*) given a Bonferroni p-value threshold = 5.68×10^-3^. (B) gsMap brain region results for (i) Essential Tremor, (ii) Parkinson’s disease, (iii) dystonia, (iv) and cognition, mapped to adult mouse cerebellum spatial transcriptomic data. Each point presents that spot’s association with trait specific GWAS signatures, with smaller Cauchy P-values in red and larger ones in blue. (C) gsMap cell type results for (i) Essential Tremor, (ii) Parkinson’s disease, (iii) dystonia, (iv) and cognition, mapped to adult mouse cerebellum spatial transcriptomic data. Each point presents that spot’s association with trait specific GWAS signatures, with smaller Cauchy P-values in red and larger ones in blue.

### ET PRS and conditioned ET PRS performances across conditions

Polygenic risk scores were calculated based on ET GWAS^6^ summary statistics and on conditioned ET GWAS (adjusted for PD and cognition) summary statistics generated from mtCOJO. The performances of these two models were evaluated at a 90^th^ percentile cut-off to see if adjusting SNP associations shared between ET and ET-Plus related phenotypes would improve patient identification among related phenotype cohorts. A McNemar test revealed that the native unadjusted ET PRS model preformed 1.33% (95% CI: [0.30% - 2.35%]; p = 0.0129) better than the conditioned ET PRS model. Given an estimated 5% disease prevalence, the accuracy of the native unadjusted PRS (accuracy = 87.93%; 95% CI: [86.93 – 88.89%]) is slightly better than the conditioned PRS (accuracy = 86.98%; 95% CI: [85.95 – 87.96%]). However, for this and all other diagnostic measures, confidence intervals were overlapping and thus differences are non-significant. The diagnostic test results for both models are shown in Figure 3.

**Figure 3.**
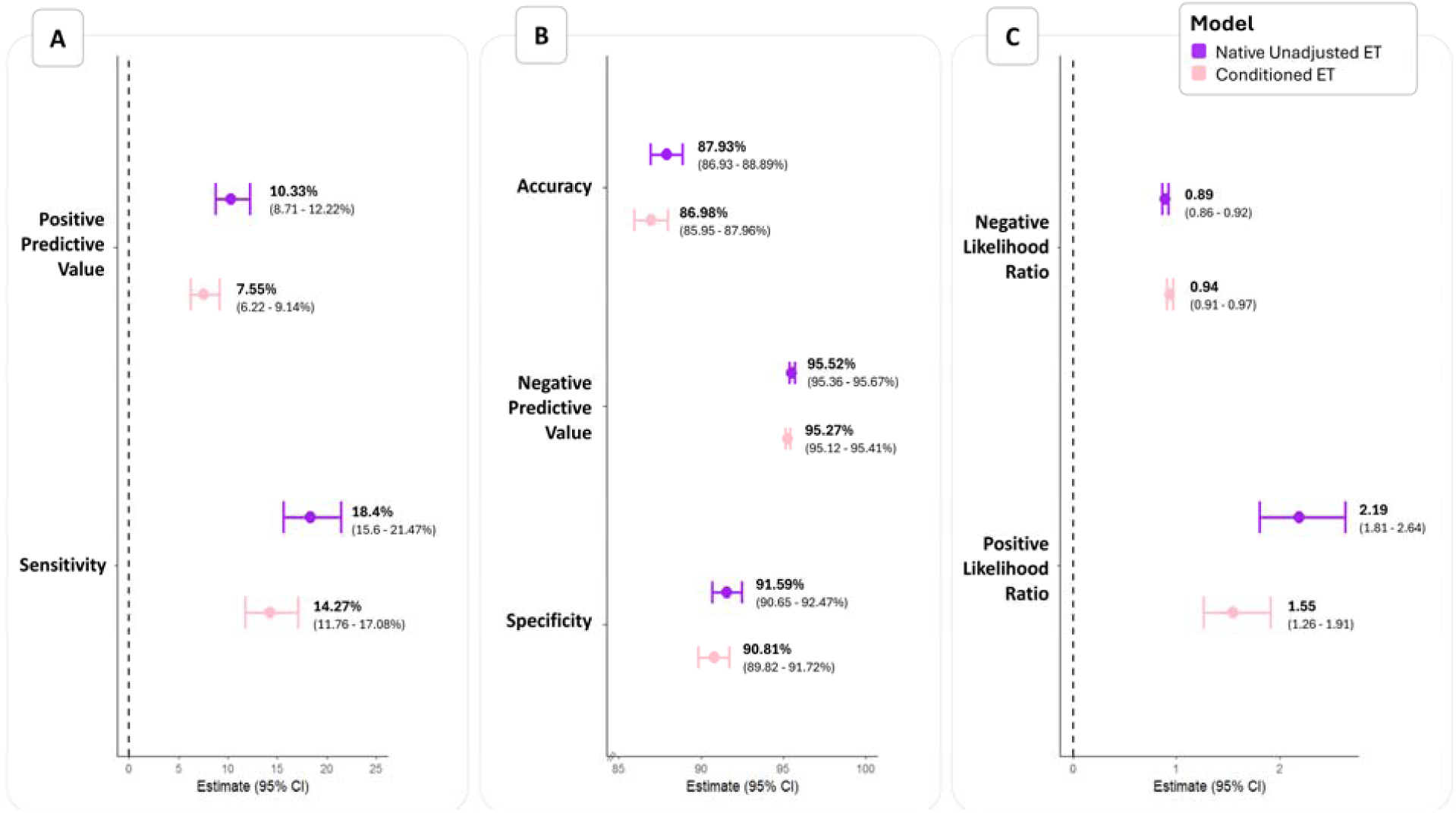
Native ET PRS model and conditioned ET PRS model performance. Model performance estimates from the diagnostic tests done using the 90^th^ percentile as a threshold where samples with PRS above the threshold test positive for ET and those below test negative. Results for the native PRS model are shown in purple, while results for the conditioned PRS model are shown in pink. Accuracy, positive predictive value, negative predictive value, sensitivity, and specificity as reported a percentages. 95% confidence intervals are reported for all tests is brackets. (A) Positive predictive value and sensitivity are reported together since they are proximal on the percentage scale. (B) Accuracy, negative predictive value, and specificity are reported together since they are proximal as percentages. (C) Positive likelihood ratio and negative likelihood ratio are reported together since they are reported a ratios.

## Discussion

ET is the most common movement disorder globally, and yet there exists controversy around a consensus definition of the condition. Across affected individuals, ET can present as a spectrum of phenotypes, where tremor affects different regions of the upper extremities at different levels of severity between individuals.^24^ Further complicating things, ET patients may also present with soft signs of other neurological disorders.^3^ The clinical heterogeneity is a challenge for ET diagnosis and study which prompted us to examine what makes Pure-ET unique from other related conditions through a genetic lens.^24^

In this work, we examined ET and the ET-Plus related phenotypes of PD, dystonia, and cognition, to identify how common variant associations across the brain differ by phenotype and to explore if conditioning ET on PD and cognition could improve PRS diagnostics. In our spatial transcriptomic analysis across the whole mouse brain and the mouse cerebellum, we observed overlapping common variant-region and common variant-cell type associations across phenotypes. No one region nor one cell type was uniquely associated to ET. However, it is encouraging that the most significant association found across the cerebellum for ET GWAS signal mapping was in oligodendrocytes which have been found to be relevant in ET from single cell ET patient data.^25^ Additionally, looking at the GWAS association mappings on the brain, there appears to be some subtle differences in the most strongly associated regions between each phenotype’s map when resolved at a higher resolution, but there are also many regions and cell types with shared GWAS association mapping.

For the PRS model performances across ET, PD, dystonia, cerebellar ataxia, cognitively impaired, and healthy samples using a 90^th^ percentile cut-off, the conditioned GWAS (conditioned for PD and cognition) informed PRS performed 1.33% (95% CI: [0.30% - 2.35%]; p = 0.0129) worse than the native unadjusted ET GWAS informed PRS. The diagnostic performance metrics appeared slightly better for the native ET GWAS model, however, confidence intervals were overlapping, so these differences are not significant. Overall, performance differences across the models are negligibly different, so we cannot conclude whether shared genetic risk between ET, PD, and cognition help to define ET or exists as noise that obscure unique genetic risk signatures for ET. Despite this, what can be noted is that ET PRS are still not clinically useful given the better performing model had a positive predictive value of only 10.33% [8.71% - 12.22%], meaning that for every positive test result only ∼1/10 are truly ET patients.

Taken together, this evidence suggests that we do not have a strong ability to decern ET from related movement disorders or cognitive phenotypes using existing common variant disease associations. Perhaps looking are rare variants or other less explored variant types in ET is a logical next step to better understand the unique genetic risk factors that set ET apart from other disorders. However, another course of action would be to further challenge the classification of ET moving forward in genetic studies. Perhaps if we considered ET as a spectrum of disease or conditions with distinct subtypes for genetic analysis, we could yield improved findings. We know that ET has a strong genetic basis and yet, for decades we have struggled to identify replicable genetic risk factors which drive ET.^26^ When patients in genetic studies are classified on a binary of ET and unaffected, we may end up grouping samples together of vastly different ET subtypes, which may create noise that obscures the genetic drivers of ET. Rather, it may instead be beneficial to sub-divide ET patients on a more refined scale not just capturing Pure-ET or ET-Plus, but to also consider the precise regions affected, tremor context (kinetic, postural, isometric), and the severity of tremor, to more finely subdivide ET, as proposed by Fanning *et. al*.^24^ The Essential Tremor Rating Assessment Scale performance subscale (TETRAS-PS) is a measurement tool which can be used to de-binarize ET for future genetic studies, by providing improved power and nuance though a continuous variable that represents tremor severity.^24^ This scoring paired with grouping ET patients based on the precise regions affected by tremor as well as additional motor and non-motor symptoms may indeed help to better resolve unique genetic underpinnings of ET. In other words, by subdividing ET into more precise categories and thus recognizing inter-group variability in ET, we may reduce intra-group variability to sharpen biological signals to benefit genetic research.

Ultimately, we could not resolve a unique genetic character to ET based on our current SNP-disease association knowledge. The heterogeneity in ET is a consistent challenge in ET research, which may obscure important ET genetic associations. Rather than dismissing this heterogeneity as noise, perhaps we should leverage it instead to more finely subdivide ET, even going beyond the ET-Plus concept. Reconceptualizing ET as a family of related disorders with distinct sub-groups may remedy some of the heterogeneous challenges of ET. Such a framework may allow us to more precisely study, diagnose, and ultimately treat ET.

## Supporting information

esupp

## Data Availability

Mouse whole brain spatial data was specifically downloaded from https://ftp.cngb.org/pub/SciRAID/stomics/STDS0000058/stomics/ (Mouse_brain.h5ad) with single cell annotations transferred from (Mouse_brain_cell_bin.h5ad) found at the same location. The annotation transfer is performed through scVI integration as described in the scvi-tools tutorial: https://docs.scvi-tools.org/en/stable/tutorials/notebooks/scrna/tabula_muris.html.

Mouse cerebellum spatial data was specifically downloaded from https://db.cngb.org/stomics/cbmsta/download/ (Mouse1_T175.h5ad).

gsMap resource bundle: https://yanglab.westlake.edu.cn/data/gsMap/gsMap_resource.tar.gz

This study used data from the *All of Us* Research Program’s Controlled Tier Dataset version 8, available to authorized users on the <u>Researcher Workbench</u>.

## Acknowledgements

We gratefully acknowledge *All of Us* participants for their contributions, without whom this research would not have been possible. We also thank the National Institutes of Health’s *<u>All of</u> <u>Us</u>* <u>Research Program</u> for making available the participant data examined in this study.

## Financial disclosures

M.M. received a doctoral student fellowship from the Canadian Institutes of Health Research (CIHR) (FRN193300).

## Notes

### Competing Interest Statement

The authors have declared no competing interest.

### Funding Statement

The Rouleau laboratory had support from the Canadian Institutes of Health Research (CIHR) Foundation Award. M.M. received a doctoral student fellowship from the Canadian Institutes of Health Research (CIHR) (FRN193300).

### Author Declarations

The McGill University Health Centre Research Ethics Board approved this work (Reference number: IRB00010120)

