## Supplementary material for "Insights into Essential Tremor and Essential Tremor-Plus from Common Variants": esupp

### Data Acquisition

#### Summary Statistics

Genome-wide association study (GWAS) summary statistics for Essential Tremor (ET), Parkinson's Disease (PD), Dystonia, and Cognition used for gsMap and mtCOJO conditioning are detailed in Table S1.

**Table S1: Origin of GWAS summary statistics used in analyses**

| Phenotype | Article | Number of Cases | Number of Controls | Ancestry | Publication Year |
|---|---|---|---|---|---|
| Essential Tremor | GWAS meta-analysis reveals key risk loci in essential tremor pathogenesis[1] | 16,480 | 1,936,173 | European | 2024 |
| Parkinson's Disease | Identification of novel risk loci, causal insights, and heritable risk for Parkinson's disease: a meta-analysis of genome-wide association studies[2] | 37,688 | 1,417,791 | European | 2019 |
| Dystonia | Genetic Risk Factors in Isolated Dystonia Escape Genome-Wide Association Studies[3] | 4,303 | 2,362 | European | 2024 |
| Cognition | Cognitive processing speed and accuracy are intrinsically different in genetic architecture and brain phenotypes[4] | 2,266,733 [Continuous trait] | | European | 2024 |

#### Polygenic Risk Score Cohort

Polygenic risk score (PRS) were calculated from native unadjusted ET GWAS summary statistics as well as the summary statistics from the ET GWAS conditioned on PD and cognition. The cohort after sample level quality control that these PRS models were applied to is described in full detail in Table S2.

**Table S2: Description of cohort that PRS was applied to.**

| Phenotype | Age (Mean [SD] ) | Sex (% Female) | Cohort Size |
|---|---|---|---|
| Essential Tremor | 77.36 [6.41] | 49.22% | 701 |
| Parkinson's Disease | 78.34 [6.77] | 35.67% | 869 |
| Dystonia | 75.47 [6.22] | 65.40% | 607 |
| Cerebellar Ataxia | 77.12 [7.42] | 48.43% | 64 |
| Cognitive Impairment | 77.8 [6.99] | 54.12% | 742 |
| Healthy Individuals | 75.73 [6.12] | 52.48% | 1394 |

### gsMap GWAS signal spatial mapping

Spatially resolved maps of ET, PD, dystonia, and cognition associated cells derived through GWAS and spatial transcriptomic data were obtained through gsMap. Across the adult mouse whole brain and cerebellum, the spatial regions and cell types significantly associated to a given trait following multiple hypothesis correction as determined by Cauchy combination is shown in Table S3

**Table S3: All gsMap significant associations across phenotypes for mouse whole brain and cerebellum**

| *Spatial Data* | *Phenotype* | *Annotation* | *P Cauchy* |
|---|---|---|---|
| *Adult Mouse Brain* | Cognition | Cortex_L6 | 1.33E-18 |
| *Adult Mouse Brain* | Cognition | Subiculum | 2.73E-17 |
| *Adult Mouse Brain* | Cognition | Cortex-L2/3 | 1.83E-16 |
| *Adult Mouse Brain* | Cognition | Stratum lacunosum/raditum of CA1 | 1.99E-16 |
| *Adult Mouse Brain* | Cognition | Cortex_L5 | 2.17E-16 |
| *Adult Mouse Brain* | Cognition | Cortex_L4 | 2.67E-16 |
| *Adult Mouse Brain* | Cognition | Stratum oriens of CA1 | 6.66E-16 |
| *Adult Mouse Brain* | Cognition | Dentate gyrus | 2.11E-15 |
| *Adult Mouse Brain* | Cognition | EX L5/6 | 2.44E-15 |
| *Adult Mouse Brain* | Cognition | Lateral-ventral cortex | 6.88E-15 |
| *Adult Mouse Brain* | Cognition | Thalamus | 3.87E-14 |
| *Adult Mouse Brain* | Cognition | Posterior amygdalar nucleus | 7.83E-14 |
| *Adult Mouse Brain* | Cognition | Hippocampus cornu ammonis area 1 (CA1) | 8.38E-14 |

| | | | |
|---|---|---|---|
| *Adult Mouse Brain* | Cognition | Molecular layer of dentate gyrus | 8.65E-14 |
| *Adult Mouse Brain* | Cognition | EX L4 | 7.16E-13 |
| *Adult Mouse Brain* | Cognition | EX L2/3 | 2.53E-12 |
| *Adult Mouse Brain* | Cognition | Cortical amygdalar area | 7.03E-12 |
| *Adult Mouse Brain* | Cognition | EX CA | 3.84E-11 |
| *Adult Mouse Cerebellum* | Cognition | Golgi | 1.47E-10 |
| *Adult Mouse Cerebellum* | Cognition | ODC | 2.04E-10 |
| *Adult Mouse Cerebellum* | Cognition | granular layer | 3.80E-10 |
| *Adult Mouse Cerebellum* | Cognition | UBC | 3.84E-10 |
| *Adult Mouse Cerebellum* | Cognition | Granule | 4.61E-10 |
| *Adult Mouse Brain* | Cognition | EX | 5.34E-10 |
| *Adult Mouse Cerebellum* | Cognition | OPC | 1.89E-09 |
| *Adult Mouse Brain* | Cognition | IN Pvalb+ | 1.55E-08 |
| *Adult Mouse Cerebellum* | Cognition | purkinje layer | 2.37E-08 |
| *Adult Mouse Cerebellum* | Cognition | white matter | 2.47E-08 |
| *Adult Mouse Brain* | Cognition | GN DG | 3.33E-08 |
| *Adult Mouse Cerebellum* | Cognition | Astrocyte | 3.74E-08 |
| *Adult Mouse Cerebellum* | Cognition | Purkinje | 4.67E-08 |
| *Adult Mouse Cerebellum* | Cognition | Endothelial_stalk | 4.96E-08 |
| *Adult Mouse Cerebellum* | Cognition | Endothelial_mural | 6.46E-08 |
| *Adult Mouse Cerebellum* | Cognition | Ependymal | 7.01E-08 |
| *Adult Mouse Cerebellum* | Cognition | Microglia | 7.07E-08 |
| *Adult Mouse Cerebellum* | Cognition | Bergmann | 7.63E-08 |
| *Adult Mouse Cerebellum* | Cognition | PLI | 1.72E-07 |
| *Adult Mouse Brain* | Cognition | Fiber tract | 1.84E-07 |
| *Adult Mouse Cerebellum* | Cognition | Choroid | 2.96E-07 |
| *Adult Mouse Brain* | Cognition | IN Sst+ | 7.13E-07 |
| *Adult Mouse Cerebellum* | Parkinson's Disease | Bergmann | 1.07E-06 |
| *Adult Mouse Cerebellum* | Cognition | molecular layer | 1.11E-06 |
| *Adult Mouse Brain* | Cognition | Astr1 | 1.29E-06 |
| *Adult Mouse Brain* | Cognition | Astr2 | 1.44E-06 |
| *Adult Mouse Cerebellum* | Parkinson's Disease | purkinje layer | 1.56E-06 |
| *Adult Mouse Brain* | Cognition | Olig | 2.53E-06 |
| *Adult Mouse Brain* | Cognition | Meninges | 3.43E-06 |
| *Adult Mouse Brain* | Cognition | Unknown | 3.67E-06 |
| ***Adult Mouse Brain*** | **Essential Tremor** | **Cortex_L5** | **4.11E-06** |
| *Adult Mouse Cerebellum* | Parkinson's Disease | Purkinje | 6.98E-06 |
| *Adult Mouse Brain* | Cognition | Astr4 | 7.64E-06 |

| | | | |
|---|---|---|---|
| *Adult Mouse Cerebellum* | Cognition | MLI1 | 7.92E-06 |
| ***Adult Mouse Cerebellum*** | **Essential Tremor** | **ODC** | **1.55E-05** |
| *Adult Mouse Brain* | Parkinson's Disease | Substandia nigra/Ventral tegmental area | 2.09E-05 |
| ***Adult Mouse Brain*** | **Essential Tremor** | **Subiculum** | **2.87E-05** |
| *Adult Mouse Brain* | Cognition | Midbrain | 5.71E-05 |
| ***Adult Mouse Cerebellum*** | **Essential Tremor** | **purkinje layer** | **6.47E-05** |
| *Adult Mouse Brain* | Cognition | EX L6 | 6.54E-05 |
| *Adult Mouse Cerebellum* | Parkinson's Disease | Golgi | 7.44E-05 |
| *Adult Mouse Cerebellum* | Cognition | Macrophage | 8.70E-05 |
| ***Adult Mouse Cerebellum*** | **Essential Tremor** | **Golgi** | **9.63E-05** |
| ***Adult Mouse Cerebellum*** | **Essential Tremor** | **Bergmann** | **9.84E-05** |
| ***Adult Mouse Brain*** | **Essential Tremor** | **CA1** | **1.23E-04** |
| ***Adult Mouse Cerebellum*** | **Essential Tremor** | **Purkinje** | **1.42E-04** |
| ***Adult Mouse Cerebellum*** | **Essential Tremor** | **granular layer** | **1.45E-04** |
| ***Adult Mouse Brain*** | **Essential Tremor** | **Cortex_L6** | **1.71E-04** |
| ***Adult Mouse Cerebellum*** | **Essential Tremor** | **UBC** | **2.21E-04** |
| *Adult Mouse Cerebellum* | Parkinson's Disease | Ependymal | 2.35E-04 |
| *Adult Mouse Brain* | Cognition | Substandia nigra/Ventral tegmental area | 2.54E-04 |
| ***Adult Mouse Cerebellum*** | **Essential Tremor** | **Granule** | **2.92E-04** |
| *Adult Mouse Cerebellum* | Parkinson's Disease | granular layer | 3.63E-04 |
| *Adult Mouse Cerebellum* | Parkinson's Disease | Choroid | 4.87E-04 |
| *Adult Mouse Cerebellum* | Parkinson's Disease | UBC | 5.25E-04 |
| *Adult Mouse Cerebellum* | Parkinson's Disease | Granule | 6.41E-04 |
| *Adult Mouse Cerebellum* | Parkinson's Disease | Astrocyte | 7.02E-04 |
| *Adult Mouse Cerebellum* | Parkinson's Disease | Endothelial_stalk | 7.14E-04 |
| ***Adult Mouse Cerebellum*** | **Essential Tremor** | **Astrocyte** | **8.36E-04** |
| *Adult Mouse Cerebellum* | Cognition | MLI2 | 8.53E-04 |
| ***Adult Mouse Cerebellum*** | **Essential Tremor** | **Endothelial_mural** | **8.53E-04** |
| *Adult Mouse Cerebellum* | Parkinson's Disease | PLI | 8.70E-04 |
| ***Adult Mouse Cerebellum*** | **Essential Tremor** | **Ependymal** | **9.01E-04** |
| ***Adult Mouse Cerebellum*** | **Essential Tremor** | **Choroid** | **1.02E-03** |
| *Adult Mouse Cerebellum* | Parkinson's Disease | Endothelial_mural | 1.51E-03 |
| *Adult Mouse Cerebellum* | Parkinson's Disease | molecular layer | 1.53E-03 |
| ***Adult Mouse Cerebellum*** | **Essential Tremor** | **OPC** | **1.81E-03** |
| *Adult Mouse Cerebellum* | Parkinson's Disease | ODC | 2.16E-03 |
| ***Adult Mouse Cerebellum*** | **Essential Tremor** | **PLI** | **2.41E-03** |
| ***Adult Mouse Cerebellum*** | **Essential Tremor** | **molecular layer** | **2.53E-03** |

| | | | |
|---|---|---|---|
| ***Adult Mouse Cerebellum*** | **Essential Tremor** | **Microglia** | **2.59E-03** |
| ***Adult Mouse Cerebellum*** | **Essential Tremor** | **white matter** | **3.57E-03** |

*All Essential Tremor significant associations are shown in bold. Spatial-Phenotype associations are presented from smallest to largest p-Cauchy.

### Essential Tremor GWAS conditioned on Parkinson's Disease and Cognition

Using mtCOJO, ET GWAS[1] summary statistics were conditioned on PD[2] and cognition[4] GWAS summary statistics. The resulting conditioned GWAS summary statistic SNP associations were visualized as a Manhattan plot in R using ggplot. This Manhattan plot is depicted in Figure S1.

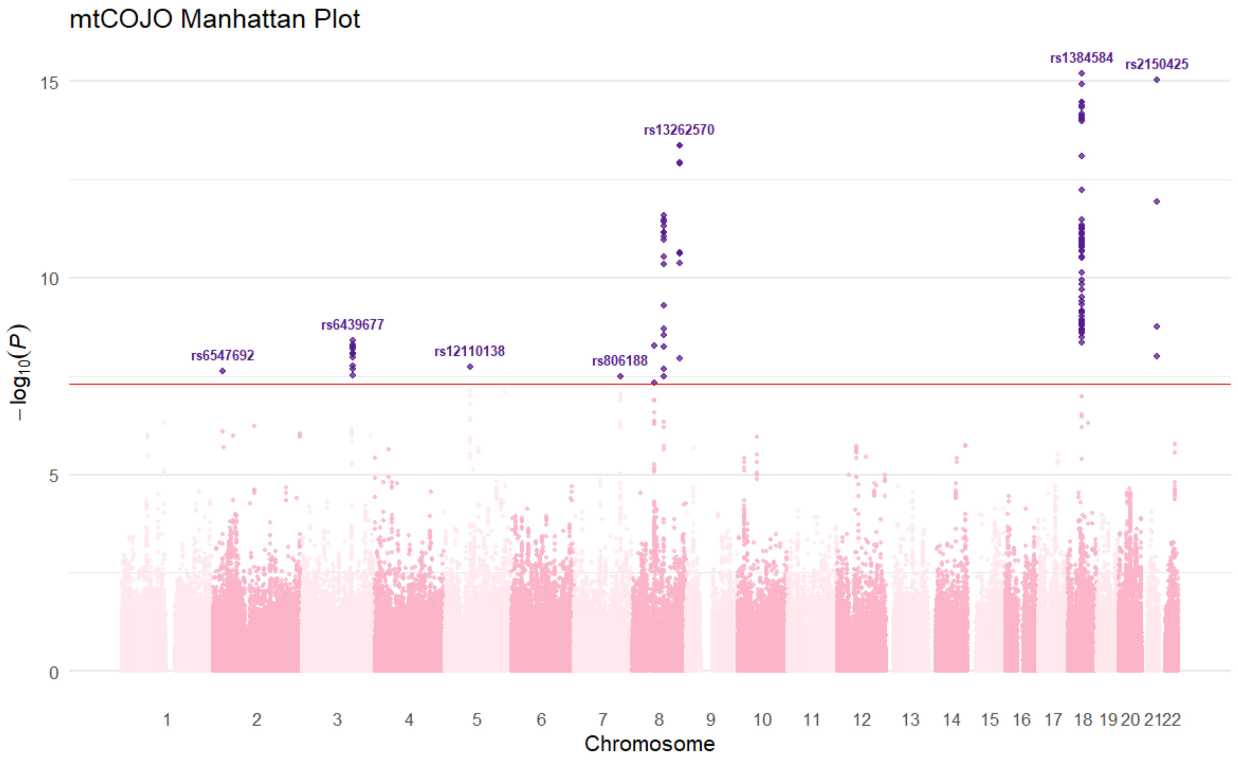


**Figure S1: Manhattan plot of ET GWAS conditioned on Parkinson's Disease and cognition**. Adjusted ET-SNP associations are shown in the Manhattan plot by chromosome after conditioning on Parkinson's Disease and Cognition GWAS summary statistics using mtCOJO. Significant SNPs associations of the conditioned GWAS are shown in purple.

### Code

##### Mapping Single Cell Annotations to Whole Mouse Brain Spatial Transcriptomics Data

```
import scvi
import scanpy as sc
import anndata as ad

# get data:
adata = sc.read_h5ad("/home/medeiros/scratch/spatial/spatial_data/Mouse_brain.h5ad")
adata_bin = sc.read_h5ad("/home/medeiros/scratch/spatial/spatial_data/Mouse_brain_cel
l_bin.h5ad")

# Set adata.X to counts
adata.X = adata.layers["count"]
adata_bin.X = adata_bin.layers["counts"]

# Add a batch/dataset label so SCVI can distinguish them
adata.obs["dataset"] = "original"
adata_bin.obs["dataset"] = "binned"

# Make sure var_names match correctly by gene name
adata.var_names = adata.var.index
adata_bin.var_names = adata_bin.var["Gene"]

# Align genes between the two datasets
adata, adata_bin = adata[:, adata.var_names.isin(adata_bin.var_names)], adata_bin[:,
adata_bin.var_names.isin(adata.var_names)]

# Reorder gene columns to match
adata_bin = adata_bin[:, adata.var_names]

# Concatenate into one AnnData object
adata_combined = ad.concat([adata, adata_bin])


# Integration with scVI (https://docs.scvi-tools.org/en/stable/tutorials/notebooks/sc
rna/tabula_muris.html)
# Setup scvi
scvi.model.SCVI.setup_anndata(adata_combined, batch_key="dataset")

# Train the model
model = scvi.model.SCVI(adata_combined)
model.train()

# Obtain the latent representations
SCVI_LATENT_KEY = "X_scVI"
adata_combined.obsm[SCVI_LATENT_KEY] = model.get_latent_representation()

# save
adata_combined.write("/home/medeiros/scratch/spatial/spatial_data/Mouse_brain_with_bi
n_annotations.h5ad")
```

#### gsMap Pipeline

Run for whole mouse brain and mouse cerebellum separately:

##### Step 1: Run latent Representations

```
gsmap run_find_latent_representations \
    --workdir '/home/medeiros/scratch/spatial/MOUSE_BRAIN_wrk' \
    --sample_name 'MOUSE_ET' \
    --input_hdf5_path '/home/medeiros/scratch/spatial/spatial_data/Mouse_brain_with_b
in_annotations_edited.h5ad' \
    --annotation 'annotation' \
    --data_layer 'count'
```

##### Step 2: Run latent to gene

```
gsmap run_latent_to_gene \
    --workdir '/home/medeiros/scratch/spatial/MOUSE_BRAIN_wrk' \
    --sample_name 'MOUSE_ET' \
    --annotation 'annotation' \
    --latent_representation 'latent_GVAE' \
    --num_neighbour 51 \
    --num_neighbour_spatial 201 \
    --homolog_file '/home/medeiros/scratch/spatial/test_data/gsMap_resource/homologs/
mouse_human_homologs.txt'
```

##### Step 3: Run Generate LD Score

```
CHROM=$1

gsmap run_generate_ldscore \
        --workdir '/home/medeiros/scratch/spatial/MOUSE_BRAIN_wrk' \
        --sample_name 'MOUSE_ET' \
        --chrom $CHROM \
        --bfile_root '/home/medeiros/scratch/spatial/test_data/gsMap_resource/LD_Refe
rence_Panel/1000G_EUR_Phase3_plink/1000G.EUR.QC' \
        --keep_snp_root '/home/medeiros/scratch/spatial/test_data/gsMap_resource/LDSC
_resource/hapmap3_snps/hm' \
        --gtf_annotation_file '/home/medeiros/scratch/spatial/test_data/gsMap_resourc
e/genome_annotation/gtf/gencode.v46lift37.basic.annotation.gtf' \
        --gene_window_size 50000 \
        --enhancer_annotation_file '/home/medeiros/scratch/spatial/test_data/gsMap_re
source/genome_annotation/enhancer/by_tissue/ALL/ABC_roadmap_merged.bed' \
        --snp_multiple_enhancer_strategy 'max_mkscore' \
        --gene_window_enhancer_priority 'gene_window_first'
```

##### Step 4: Run Spatial LDSC

```
# Run this seperatly for each phenotype (seperate summary statistics input)

gsmap run_spatial_ldsc \
    --workdir '/home/medeiros/scratch/spatial/MOUSE_BRAIN_wrk' \
    --sample_name 'MOUSE_ET' \
    --trait_name 'ET' \
    --sumstats_file '/home/medeiros/scratch/spatial/data/sum_stats/ET_gsmap_sumstats.
```

```
sumstats.gz' \
    --w_file '/home/medeiros/scratch/spatial/test_data/gsMap_resource/LDSC_resource/w
eights_hm3_no_hla/weights.' \
    --num_processes 4
```

##### Step 5: Run Cauchy combination

```
gsmap run_cauchy_combination \
    --workdir '/home/medeiros/scratch/spatial/MOUSE_BRAIN_wrk' \
    --sample_name 'MOUSE_ET'\
    --trait_name 'ET' \
    --annotation 'annotation'
```

##### Step 6: Run Report

```
gsmap run_report \
    --workdir '/home/medeiros/scratch/spatial/MOUSE_BRAIN_wrk' \
    --sample_name 'MOUSE_ET'\
    --trait_name 'ET' \
    --annotation 'annotation' \
    --sumstats_file '/home/medeiros/scratch/spatial/data/sum_stats/ET_gsmap_sumstats.
sumstats.gz' \
    --top_corr_genes 50
```

#### Conditioning ET GWAS on PD and cognition

```
gcta64 --mbfile mtcojo_ref_data.txt --mtcojo-file mtCOJO_summary_data_noDystonia.txt
--ref-ld-chr /home/medeiros/scratch/spatial/data/sum_stats/mtCOJO/1KG/eur_w_ld_chr/ -
-w-ld-chr /home/medeiros/scratch/spatial/data/sum_stats/mtCOJO/1KG/eur_w_ld_chr/ --ou
t test_mtcojo_result
```

Contents of input files:

```
head mtCOJO_summary_data_noDystonia.txt
ET          ET_sum_stats_clean.txt.pruged         0.00844     0.01
PD          PD_sum_stats_clean.txt.pruged         0.0387       0.00572

head mtcojo_ref_data.txt
/home/medeiros/scratch/spatial/data/sum_stats/mtCOJO/1KG/1000G.EUR.QC
```

#### PRS Calculation

```
# All chromosomes were merged prior to this step.
# QC SNPs
# calculate freqs
plink --bfile european_cohort_merged  \
--memory 15000 \
--freq \
--out raw_european_cohort_merged

# determine ambiguous SNPs
```

```
awk '/^[^#]/ { if( $5>0.4 && $5<0.6 && ( ($3=="A" && $4=="T") || ($4=="T" && $3=="A")
|| ($3=="C" && $4=="G") || ($4=="G" && $3=="C") ) ) { print $0 }}' \
raw_european_cohort_merged.frq > ambiguous_SNPs_rsIDs_unrelated_european_merged.txt

# SNP QC - filtering
plink \
--bfile european_cohort_merged \
--exclude ambiguous_SNPs_rsIDs_unrelated_european_merged.txt \
--allow-no-sex \
--geno 0.01 \
--hwe 0.000001 \
--maf 0.01 \
--make-bed \
--memory 79000 \
--out cleaned_rsIDs_unrelated_european_merged

# Run PRS-CS
python PRScs/PRScs.py \
--ref_dir=ldblk_1kg_eur \
--bim_prefix=cleaned_rsIDs_unrelated_european_merged \
--sst_file=cleaned_ET_sumstats.txt \
--n_gwas=1952653 \
--out_dir=PRS_output

# Clean up:
cat PRS_output_pst_eff_a1_b0.5_phiauto_chr*.txt  | sort -n -k1 > PRS_pst_eff_a1_b0.5_
phiauto_allchr.txt

# Would also cat all files together then do this:
# create the input file for PRS that includes the inferred posterior effect size for
each SNP
awk '{print $2, $4, $6}' PRS_pst_eff_a1_b0.5_phiauto_allchr.txt | sed 's/ /\t/g' > PR
S_pst_eff_a1_b0.5_phiauto_allchr.PLINKscore


# compute individual-level PRS using PLINK SCORE
plink \
--bfile cleaned_rsIDs_unrelated_european_merged \
--score PRS_pst_eff_a1_b0.5_phiauto_allchr.PLINKscore no-mean-imputation \
--out ET_PRS_pst_eff_a1_b0.5_phiauto_allchr.PLINKscore.txt

# EXTRACT PCs for SCORE Regressions:
plink --bfile cleaned_rsIDs_unrelated_european_merged \
--pca 10 \
--out cleaned_cohort_PC10.txt
```

Scores were corrected in R:

```
# fit regression model
fit <- lm(SCORE ~ PC1 + PC2 + PC3 + PC4 + PC5 + PC6 + PC7 + PC8 + PC9 + PC10, ET)

# obtain residuals:
residual_output <- fit$residuals
```

```
# correct scores
ET$CORRECTED_SCORE <- residual_output
```

The PRS workflow was then repeated with the conditioned GWAS summary statistics.
